# Estimating excess 1- year mortality from COVID-19 according to underlying conditions and age in England: a rapid analysis using NHS health records in 3.8 million adults

**DOI:** 10.1101/2020.03.22.20040287

**Authors:** Amitava Banerjee, Laura Pasea, Steve Harris, Arturo Gonzalez-Izquierdo, Ana Torralbo, Laura Shallcross, Mahdad Noursadeghi, Deenan Pillay, Christina Pagel, Wai Keong Wong, Claudia Langenberg, Bryan Williams, Spiros Denaxas, Harry Hemingway

**Affiliations:** Institute of Health Informatics, University College London, 222 Euston Road, London, UK; University College London Hospitals NHS Trust, 235 Euston Road, London, UK; Barts Health NHS Trust, The Royal London Hospital, Whitechapel Rd, London, UK; Division of Infection and Immunity, University College London, London, UK; Clinical Operational Research Unit, University College London (UCL), London, UK; MRC Epidemiology Unit, University of Cambridge, Cambridge, UK; Institute of Cardiovascular Science, University College London (UCL), London, UK; UCL Hospitals NIHR Biomedical Research Centre, London, UK; Health Data Research UK

## Abstract

**Background:** The medical, health service, societal and economic impact of the COVID-19 emergency has unknown effects on overall population mortality. Previous models of population mortality are based on death over days among infected people, nearly all of whom (to date at least) have underlying conditions. Models have not incorporated information on high risk conditions or their longer term background (pre-COVID-19) mortality. We estimated the excess number of deaths over 1 year under different COVID-19 incidence rates and differing mortality impacts.

**Methods:** Using population based linked primary and secondary care electronic health records in England (HDR UK - CALIBER), we report the prevalence of underlying conditions defined by UK Public Health England COVID-19 guidelines (16 March 2020) in 3,862,012 individuals aged ≥30 years from 1997-2017. We used previously validated phenotypes, openly available (https://caliberresearch.org/portal), for each condition using ICD-10 diagnosis, Read, procedure and medication codes. We estimated the 1-year mortality in each condition, and developed simple models of excess COVID-19-related deaths assuming relative risk (RR) of the impact of the emergency (compared to background mortality) of 1.2, 1.5 and 2.0.

**Findings:** 20.0% of the population are at risk according to current PHE guidelines, of which; 13.7% were age>70 years and 6.3% aged ≤70 years with ≥1 underlying condition (cardiovascular disease (2.3%), diabetes (2.2%), steroid therapy (1.9%), severe obesity (0.9%), chronic kidney disease (0.6%) and chronic obstructive pulmonary disease, COPD (0.5%). Multimorbidity (co-occurrence of ≥2 conditions in an individual) was common (10.1%). The 1-year mortality in the at-risk population was 4.46%, and age and underlying conditions combine to influence background risk, varying markedly across conditions (5.9% in age>70 years, 8.6% for COPD and 13.1% in those with ≥3 or more conditions). In a suppression scenario (at SARS CoV2 rates of 0.001% of the UK population), there would be minimal excess deaths (3 and 7 excess deaths at relative risk, RR, 1.5 and 2.0 respectively). At SARS CoV2 rates of 10% of the UK population (mitigation) the model estimates the numbers of excess deaths as: 13791, 34479 and 68957 (at RR 1.2, 1.5 and 2.0 respectively). At SARS CoV2 rates of 80% in the UK population (“do-nothing”), the model estimates the number of excess deaths as 110332, 275,830 and 551,659 (at RR 1.2, 1.5 and 2.0) respectively.

**Interpretation:** We provide the public, researchers and policy makers a simple model to estimate the excess mortality over 1 year from COVID-19, based on underlying conditions at different ages. If the relative mortality impact of COVID-19 were to be about 20% (similar magnitude as the established winter vs summer mortality excess), then the excess deaths would be 0 when 1 in 100 000 (suppression), 13791 when 1 in 10 (mitigation) and 110332 when 8 in 10 are infected (“do nothing”) scenario. However, the relative impact of COVID-19 is unknown. If the emergency were to double the mortality risk, then we estimate 7, 68957 and 551,659 excess deaths in the same scenarios. These results may inform the need for more stringent suppression measures as well as efforts to target those at highest risk for a range of preventive interventions.

## Introduction

The COVID-19 pandemic may cause excess mortality over the next year in the population both because of deaths among those infected, and because people who are not infected are experiencing social and economic upheaval; meanwhile the ability of health services to provide high quality of care for both infected and uninfected patients is increasingly threatened. The net effect of mortality of this emergency on the population is thus not only a matter of modelling an infectious disease, but modelling the mortality effects of wider medical and societal changes. One way of estimating and monitoring excess mortality is to compare observed numbers of deaths with those expected based on the background (pre-COVID-19) risks of death in the population(1).

However, current models of the population mortality impact of COVID-19 are based on age-stratified death rates over days in patients infected with COVID-19 and have not incorporated clinical information from NHS health records regarding prevalence of underlying conditions, their differing background (pre-COVID-19) long term mortality risks, or the impact of differing levels of additional risk associated with COVID-19(2). There have been limited reports so far of the excess deaths beyond those expected in specific high-risk populations(3, 4). The majority of COVID-19 deaths reported so far have occurred in patients with underlying health conditions or at older ages (5-7); this situation is changing (SH personal communication) with severe infections being treated in younger COVID-19 patients without underlying conditions. Case fatality rates(CFR) for COVID-19 vary from 0.27% to 10% according to a meta-analysis(8), possibly explained by differing demography, testing strategies and prevalence of underlying conditions. However, population estimates are lacking, since cases need to be tested and most cases go untested. Moreover, since testing is more common in hospital patients, included individuals are sicker.

Currently the UK government has not implemented full suppression strategies. Thus, for example, people are still ‘socially close’ in work and public places. Social distancing and other strategies have focussed on high-risk groups, with the CDC specifying “older adults, and people who have serious chronic medical conditions such as heart disease, diabetes and lung disease”(9). The UK government published guidelines on 16 March 2020, specifying particular population subgroups at high risk from COVID-19 infection(10). Based on these guidelines, the UK has moved to the pandemic’s “delay” phase and has recommended stringent “social-distancing” measures telling people how to stay away from others(11). Today (22 March 2020) the government announced that 1.5 million ‘extremely vulnerable’ people in England with underlying conditions (including COPD and heart disease) would be asked to ‘shield’ for 12 weeks, but did not release information on how this list was chosen and others not(12).

We provide estimates of 1-year mortality by underlying conditions in order to capture the potentially profound societal insult of the epidemic. We used the CALIBER resource as an open research platform with validated, reusable definitions of several hundred underlying conditions(13, 14). This includes a sample of English data from the Clinical Practice Research Database and is linked to Hospital Episodes Statistics. For information governance and other reasons it is currently not possible for clinical researchers to access nationwide NHS data. Our objectives were: (i) to provide the research and policy community and public with parameters (prevalence, background pre-COVID-19 1-year mortality risk by age and underlying conditions) to assist modelling; and (ii) to provide initial estimates of the excess COVID-19-related deaths over a 1 year period based on differing rates of infection. It is currently not known what the overall mortality impact of this emergency, which affects infected and non-infected patients will be. The relative excess in deaths in the winter compared to the summer months in the UK and many other countries is about 20%; our model in effect applies an additional all year round winter effect (i.e. relative risk of 1.2) (15-16). However, it is possible that the relative risk varies during the course of the pandemic, and at high levels of compromise in the workforce and health system, it is likely to be higher due to inability to provide timely care. Given concerns at the clinician and system levels, we therefore modelled an increased risk of mortality associated with COVID-19(RR 1.5 and 2.0).

## Methods

### Data sources

Our study had a cohort design with prospective recording of data and follow-up. The <u>C</u>linical rese<u>A</u>rch using <u>LI</u>nked <u>B</u>espoke studies and <u>E</u>lectronic health <u>R</u>ecords (CALIBER) links patient records from different data sources: primary care (Clinical Practice Research Datalink-GOLD), hospital care (Hospital Episodes Statistics), and death registry (Office of National Statistics). A description of the phenotyping process and extensive validations of cardiovascular risk factors and endpoints have been previously published (www.caliberresearch.org/portal)(13). Data linkage was performed through UK unique individual identification data (NHS number). We used primary care records for individual patients from January 1997 to January 2017. Study approval was granted by the Independent Scientific Advisory Committee of the Medicines and Healthcare products Regulatory Agency in the UK in accordance with the Declaration of Helsinki.

### Study population

Nearly all (>99%) of the English population is registered with a general practitioner; and the data used in CALIBER has been shown to be representative of the general population of England in terms of sociodemographic characteristics and overall mortality. Eligible individuals were ≥30 years of age and were registered with a general practice between 1 January 1997 and 1 January 2017 with at least one year of follow-up data. Demographic (age, gender, index of multiple deprivation (IMD) quintiles and geographic region) and baseline characteristics were recorded(17, 18). The overall population at high risk of COVID-19 and recommended for social distancing was defined as per Public Health England guidance(10).

The risk factors and diseases were defined using Read clinical terminology systems in primary care diagnosis and the 10th version of the International Statistical Classification of Diseases (ICD-10) for hospital admissions as per previously validated CALIBER phenotypes(13, 14, 17-19). Hypertension was defined based on recorded values in primary care according to the most recent guidelines: ≥140 mmHg systolic blood pressure (or ≥150 mmHg for people aged ≥60 years without diabetes and chronic kidney disease) and/or ≥90 mmHg diastolic blood pressure(17). Diabetes was defined at baseline (including type 1, type 2, or uncertain type) on the basis of coded diagnoses recorded in CPRD or HES at or before study entry(18). Severe obesity was defined as body mass index >=40kg/m^2^. CVD was defined as the 12 most common symptomatic manifestations: chronic stable angina, unstable angina, myocardial infarction, unheralded death from coronary heart disease, heart failure, cardiac arrest/sudden coronary death, transient ischaemic attack, ischaemic stroke, intracerebral haemorrhage, subarachnoid haemorrhage, peripheral arterial disease, and abdominal aortic aneurysm, as per prior studies(18,19). Multimorbidity was defined as the co-occurrence of 2 or more conditions in an individual(20). Given the recent interest in the potential roles of angiotensin-converting enzyme inhibitors (ACEI)(21, 22) and non-steroidal inflammatory drugs (NSAIDs)(23, 24), we also estimated prevalence of use of these drugs. To assess prevalent risk factors, only included records from the year prior to baseline. Follow-up ceased at the date of death or on 1 January 2017.

### Analysis

We estimated the prevalence of each underlying condition and used Kaplan-Meier estimates of 1 year all-cause mortality in the English health records sample. We used the KM estimates in each age and number of conditions cell to estimate the number of excess deaths by age bands, and number of underlying conditions. We then modelled the excess mortality from COVID-19 for relative mortality risk associated with COVID-19 (relative risk, RR) of 1.2, 1.5 and 2.0 (25, 26), at the following infection rates in the UK population: “fully suppress” (0.0001%), “partially suppress” (1%), “mitigate” (20%) or “do nothing” (80%)(2, 27). In order to project the study estimates of excess deaths to the whole UK population, we used 2018 estimates of overall population size and mortality(28). For illustration, we applied our study estimates to United Nations Population Fund(29) estimates of population size in other countries. All analyses were performed using R (version 3.4.3).

## Results

### Prevalence of underlying conditions

We included 3,862,012 individuals aged ≥30 years: 1,957, 935 (50.7%) female, 3,331,280 (86.3%) ≤70 years (mean age 43.5 in male and females) and 530732 (13.7%) >70 years (mean age 78.1 in males and 80.2 in females) (**Suppl Table 1**).

At least 20.0% of the population aged ≥ 30 years had one or more high risk condition defined by PHE 16 March 2020 guidelines; of which 13.7% were age>70 years and 6.3% aged ≤70 years who had 1 or more underlying condition (cardiovascular disease (2.3%), diabetes (2.2%), steroid therapy (1.9%), severe obesity (0.9%), chronic kidney disease (0.6%) and chronic obstructive pulmonary disease, COPD (0.5%), chronic liver disease (0.02%), chronic neurological conditions (0.02%), splenic disorders (0.01%), immune disorders (0.001%), HIV/AIDS (0.005%)(**Figure 1**).

**Figure 1.**
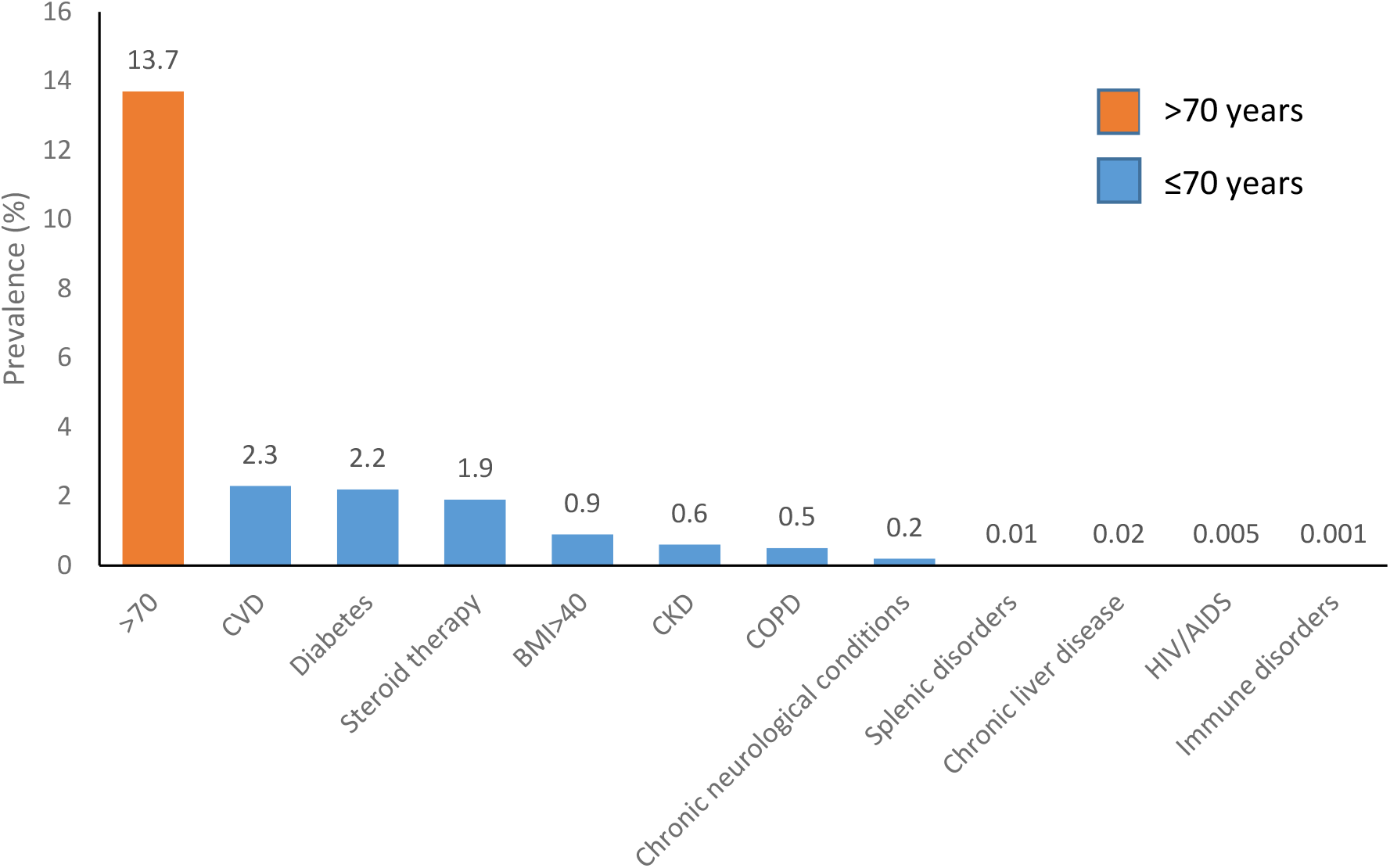
Prevalence of high-mortality risk factors for COVID-19 infection in 3,862,012 individuals.

### Multimorbidity

Multimorbidity was common (10.1%), especially in the presence of CVD, diabetes, CKD and COPD (**Suppl Table 2**). In individuals without CVD (mean age 47.5), diabetes (mean age 48.0), CKD (mean age 48.0) and COPD (mean age 48.2) respectively, 6.7%, 7.9%, 9.1%, and 9.7% still had a condition meeting criteria for social distancing. Hypertension was common: 7.4% and 31.7% in the ≤70 and >70 years subgroups respectively. In individuals aged ≤70 years, use of ACEI was not common (3.7%), compared with 14.9% in the >70 years subgroup. 14.0% of the ≤70 years population used NSAIDs, compared with 21.1% in individuals aged >70 years.

### Background (pre-COVID-19) 1-year mortality in underlying conditions

Mortality at 1-year was 4.46% among those aged >70 years or with one or more risk condition. Mortality was 6.4% (95% CI 6.2-6.5%) for CVD, 4.1% (4.0%-4.2%) for diabetes, 7.8% (7.6%-8.1%) for CKD and 8.6%(8.3-8.9%) for COPD (**Figure 2)**. Among those >70 years, one-year mortality rates for COPD, CKD, CVD and diabetes were 13.6(12.9-14.3%), 11.5%(11.0-12.1%), 10.4%(10.1-10.7%) and 8.9%(8.5-9.4%) in men and 12.3%(11.6-13.0%), 9.6%(9.3-10.0%), 10.6%(10.3-10.9%) and 9.8%(9.4-10.2%) in women.

**Figure 2.**
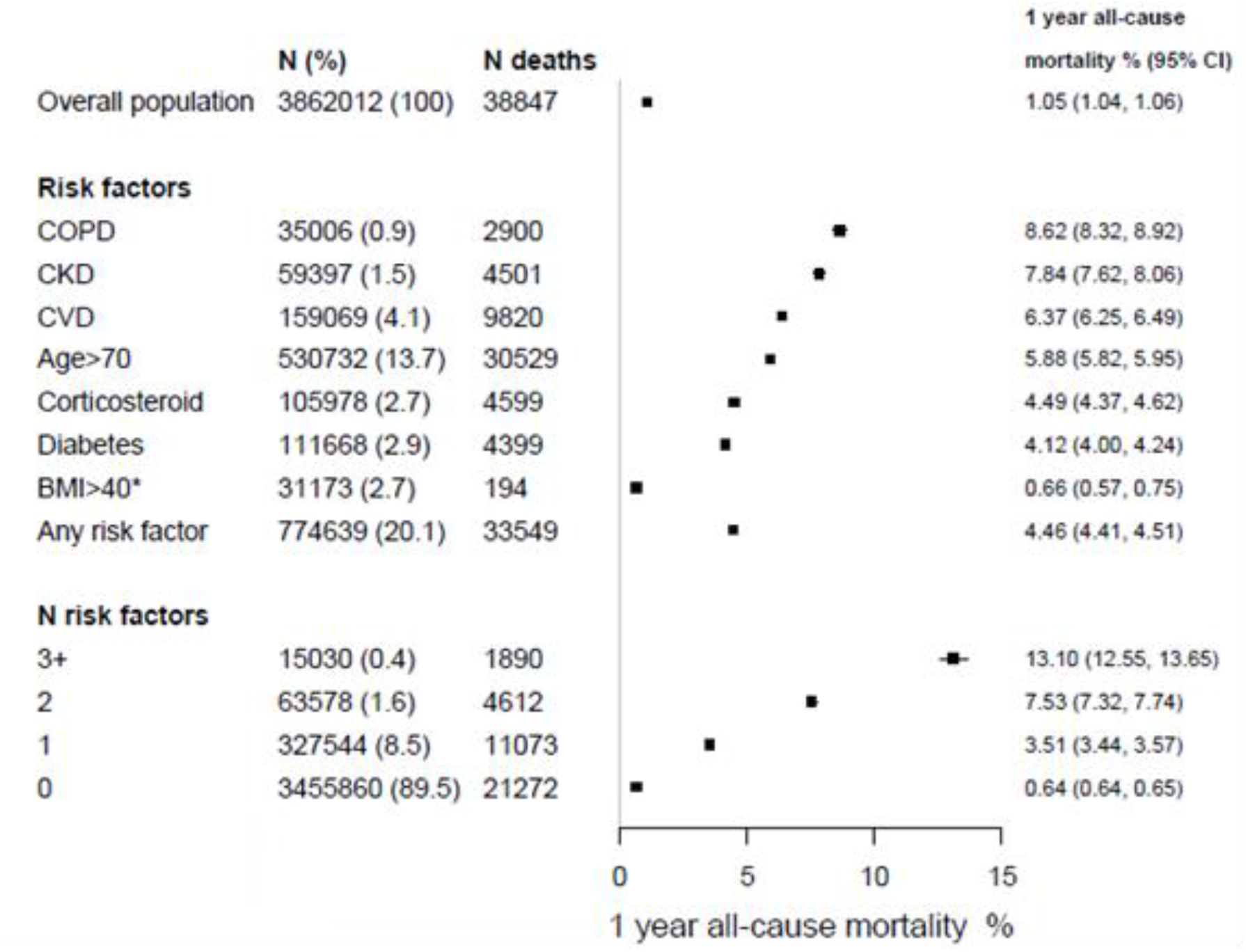
Background (pre_COVID-19) 1-year mortality in England according to underlying conditions (n=3862012)

### Background (pre-COVID-19) 1-year mortality by age and number of underlying conditions

**Figure 3** shows how age and the number of conditions combine to influence 1-year mortality risk, separately in men and women in the absence of COVID-19. For example, we found that a man aged 66-70 years of age with no underlying conditions has higher 1-year mortality than a woman aged 56-60 with one underlying condition (1.07% vs 0.91%).

**Figure 3.**
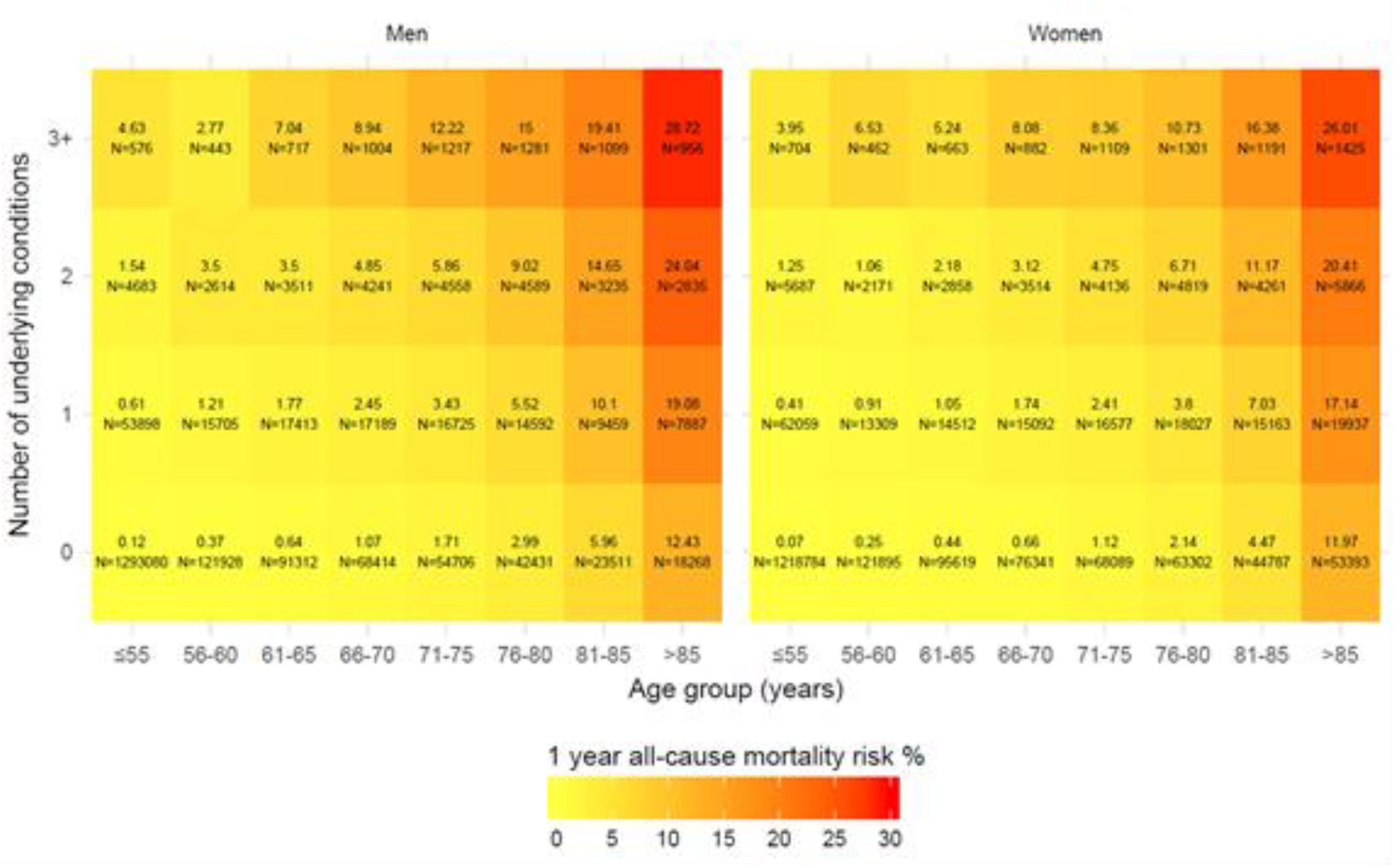
Estimated 1-year mortality according to number of underlying conditions, age and sex in England (n=3862012)

### Estimates of excess 1 year deaths in UK under different assumptions of the emergenc

At COVID-19 prevalence of 10% (mitigation) and 80%(“do-nothing”), we estimate 13791 and 110332 excess deaths at RR 1.2; 34479 and 275830 at RR 1.50 and 68957 and 551659 at RR 2.0 respectively. The corresponding deaths at “full suppression” and partial suppression rates of 0.001% and 1% respectively were 1 and 1379, 3 and 3448, and 7 and 6896. (**Figure 4**). In the USA, infection rates of 0.0001%, 20% and 80% would result in 294, 588876 and 2355503 deaths, at 2 fold increased mortality risks, respectively (**Web Table 1**).

**Figure 4.**
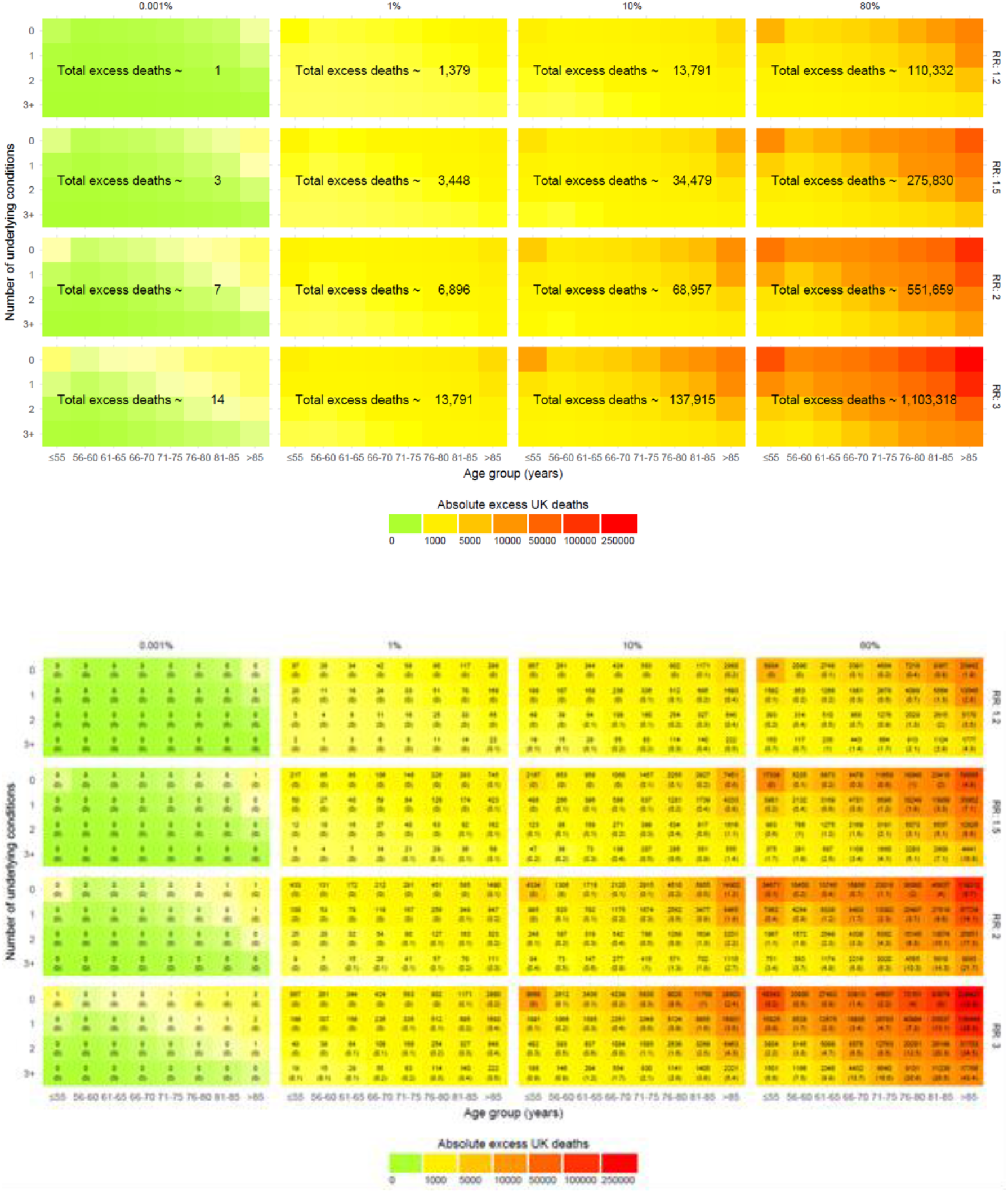
‘Estimated number of excess deaths at 1 year in the UK at different infection rates and relative risks (RR) of the impact of the current emergency.

Extrapolating from our English sample to the whole population of the UK, we estimate that 13.4 million (20% of the UK population of 66.4million) people are high risk, of whom 9.1 million >70 years of age, and 4.2 million based on having 1 or more underlying condition aged ≤70 years). Based on the background (pre-COVID-19) 1-year mortality, we observe in the CALIBER data (4.46%), there are 594423 deaths. Using these estimates gives crude mortality estimates for Italy, Germany, France, Sweden, USA and Canada, the potential numbers at high-risk were 11.9, 16.5, 13.1, 2.0, 65.8 and 7.5 million respectively(**Table 1**).

**Table 1.** Projected number of deaths in one year in high-risk groups pre-COVID-19 infection across countries.

| Country | Population (millions)<sup>1</sup> | High-risk mortality group (millions) |  |  | Baseline number of deaths in one year in high-risk group<sup>4</sup> |
| --- | --- | --- | --- | --- | --- |
|  |  | >70 years<sup>1</sup> | <70 years with underlying risk factor<sup>2</sup> | Total<sup>3</sup> |  |
| UK | 66.44 | 9.13 | 4.20 | 13.33 | 594423 |
| Italy | 59.20 | 8.11 | 3.74 | 11.88 | 529648 |
| Germany | 82.40 | 11.29 | 5.21 | 16.53 | 737213 |
| France | 65.50 | 8.97 | 4.14 | 13.14 | 586013 |
| Sweden | 10.10 | 1.38 | 0.64 | 2.03 | 90362 |
| United States of America | 329.10 | 45.09 | 20.80 | 66.02 | 2944379 |

*Assumptions based on England (CALIBER) estimates above:*
<sup>1</sup>Prevalence=13.74%
<sup>2</sup>Prevalence=6.32%
<sup>3</sup>Prevalence= 20.06%
<sup>4</sup>One year mortality (Kaplan Meier) in overall high-risk mortality group: 4.46%

## Discussion

Using a small sample of the relevant clinical data that exist in the NHS we provide an initial population-based study estimating plausible ranges of excess mortality arising from the COVID-19 pandemic in England: we provide estimates of the background (pre-COVID-19) 1-year mortality risks by underlying conditions at different ages; a major parameter that can be reliably estimated. There are implications of this rapid communication for policymakers, clinicians, and the public.

### Policy: time for stringent suppression measures

These simple estimates of excess deaths suggest that the government should (like other countries) do more in the pursuit of suppressing the epidemic whether through “enforced lockdowns” or enforced social distancing rather than voluntary measures(30).

We do not know what the relative impact of excess deaths from COVID-19 over a 1-year period will be; but the clinical concern in England over the last weeks is that ‘this is not seasonal flu’. We show that if the emergency is associated with a 20% increased mortality risk, then there will be 13791 and 110332 excess deaths in mitigation and “do nothing” scenarios and 1 death under full suppression. A doubling of the risk, although unlikely, is worth considering due to uncertainty regarding the actual relative risk associated with COVID-19 and mounting healthcare burden around the world. It would result in 68957 and 551659 excess deaths in mitigation and “do nothing” scenarios respectively. A major concern is that these relative risks may change over time: for example if the health service is unable to cope. Even in the higher RR 2.0 model, full suppression would lead to virtually no excess mortality.

### Policy: targeting ‘extremely vulnerable’ patients

Today (22 March 2020) the government announced that 1.5 million ‘extremely vulnerable’ people with underlying conditions would be asked to ‘shield’ for 12 weeks(12). The mortality risk of the conditions on this new list was not reported, nor are the national NHS data defining these conditions readily accessible for research scrutiny. It is likely that patients with conditions not on the list (as shown in figure 3) may be at as high or greater mortality than those who are; for example, those with heart failure. Cardiovascular disease is not on the list and here we show that the 1 year mortality is 6%; we found that the 1 year mortality risk of people with 2 or more conditions was 11% - but multiple morbidities co-occurring in the same individual are not on the list.

We demonstrate that on current criteria at least 20% of the population falls within the high-mortality risk category; 13.7% based on age >70 years (an arbitrary cut-off) and a further 6.3% based on having one or more underlying condition: most frequently CVD, diabetes, steroid therapy and COPD; 11.7% had 2 or more underlying conditions (multimorbidity). We show how policy might consider age in combination with underlying conditions. For example a man age 66-70 years of age with no underlying conditions is currently not considered at high risk, but we demonstrate that his background 1-year mortality (1.07%) is higher than that of a woman aged 56-60 with one underlying condition who is considered high-risk (0.91%).

### Clinicians and epidemic research

As clinical academics are re-directed to provide full-time clinical service, we believe it is important to find new, efficient clinicians to pose and answer policy relevant questions using NHS data. The team of clinicians presenting this rapid communication are in the midst of delivering a rapidly changing clinical service to patients in the COVID-19 pandemic. The motivation was to quickly provide the government, the public, researchers, and clinicians clinical information on background mortality risks (pre-COVID-19) in order to begin to estimate excess deaths in high-risk populations to inform decision-making. We encourage the community to share clinical definitions in the open HDR UK CALIBER portal and rapidly accelerate use of the NHS clinical information.

### Clinicians, healthcare workers and service

Primary care and secondary care healthcare workers have an important role in optimising guideline-recommended management of underlying conditions to lower the background risk. Missed opportunities for effective (secondary) preventive interventions are common in many countries in the management of non-communicable diseases, particularly CVDs, COPD and diabetes(31). The concern is that with COVID-19 pressures these practical actions for clinicians now will not be addressed, and chronic disease management care might even deteriorate.

### Legislation to mobilise NHS data for research

Research is a key part of the COVID-19 response, and we believe that the government should act, with legislation and other means, to massively mobilise publicly accountable access to nationwide NHS health and social care data across a large number of data custodians for researchers, clinician and policy makers NHS data across the whole country. Currently researchers can access only small (here in 5% of the UK population) samples, in which linkage to even simple data on hospital admissions requires multiple approvals, and can take years. Currently it is not possible to access NHS data on the COVID-19 epidemic as a nation-wide whole (it is fragmented by primary and secondary care, and fragmented across Scotland, Northern Ireland, Wales and England). Currently researchers cannot access detailed data across hospitals; there are small networks for critical care(32), and coronary disease(33), but no national efforts to stream real time, actionable data on the COVID-19 care and (non-fatal) outcomes. With government direction the NHS has an opportunity to create a learning health system around COVID-19, in which high-quality clinical information on COVID-19 patients (and non-infected patients) is collected: this would need to include measures of disease severity (often absent).

### Mobilising data cross government departments

Understanding how to mitigate the epidemic will require accessing and linking data about causes and consequences of this ‘insult’ across sectors including education, economy, transport. This is required in order to rapidly learn how to effectively respond to the COVID-19 epidemic. Social distancing policies may themselves influence the background mortality risks, as people may be less likely to access health and social care, and because social isolation has in many previous studies been shown to be associated with the onset and progression of cardiovascular and other diseases(34). Thus it is possible that we may have under-estimated the true 1-year mortality risks. It is not known whether sudden and systematic social distancing(35, 36), particularly if prolonged over months, has different or additional health consequences, compared to those reported from earlier studies of ‘social isolation.

### The public

We believe, has a right to see and interact with rapidly emerging research knowledge relevant to the epidemic. We provide readily understandable estimates of background (pre-COVID) 1-year risk of death according to number of underlying conditions, by age and sex. This provides context for the daily reports of the numbers of deaths (the ‘numerator’); at the time of writing, there are 177 deaths from coronavirus in the UK. It is not widely appreciated that on average 1400 people die every day in the UK(37). Of all those people dying within 1 year, it is likely that COVID-19 ‘brings forward’ the death earlier in the year. In other words there are competing causes for the mortality. The public may not be used to seeing ‘heat maps’ of mortality risk which sets the numerator in the context of the population at risk, the denominator – but this may change.

#### International harmonisation of clinical data from electronic health records

International efforts are required to harmonise definitions of underlying conditions, and the clinical syndrome and progression of COVID-19 infection, using clinically collected data in EHRs. This is fundamental for the efficient generation of reproducible knowledge to address the pandemic. To date there have been limited efforts across national or other jurisdictions and it has previously been shown that clinical data are combined in more than 60 different ways (each research group doing their own thing) to define one disease, asthma(38). The CALIBER portal (www.caliberresearch.com) is the Health Data Research UK open platform for depositing EHR phenotypes (code lists, logic, validations, use in publications)(13).

#### Illustration in US and other countries

Countries differ in the availability of population-based longitudinal EHRs to estimate prevalence and mortality among underlying conditions. The age structure and age-specific prevalence of underlying conditions and their mortality also differs between countries(39). Nonetheless our estimates might be a relevant starting point for other countries; as an illustration we show estimates of excess deaths using the background mortality from the UK.

#### Limitations and further research

This rapid communication was developed over a 72-hour period prior to posting the results on 22 March 2020; and will be peer reviewed in due course. We seek to work with others to report further estimates of prevalence and mortality for cancer, pregnancy and chemotherapy, cause-specific mortality and cause specific hospital admissions (particularly respiratory) and critical care admissions, as well as the further conditions listed today (22 March 2020) (organ transplants, cystic fibrosis, specific cancers of the blood or bone marrow and immunosuppression(12). There are many avenues for further modelling to better understand how to target different preventive interventions. An (incomplete) list includes statistical aspects (dynamic models, weekly rates of mortality, competing risks), public health aspects (regional variation, social deprivation and ethnicity), clinical aspects (including hospital and critical care admissions) and health service factors (including data on healthcare workers, and operational features). We have assumed that the impact (RR) of COVID-19 on excess mortality is the same across all risk groups, irrespective of how they are combined, which may not be the case. However, we researchers can use the estimate that we provide to allow modelling of such interactions in different subgroups within populations. We prioritised sharing estimates of 1-year mortality at a time when all clinical academics in the UK and around the globe are being called to the frontline(40). In these times, our science must cross disciplines, disease specialisms and national and jurisdictional borders even more than before and we invite colleagues to improve, develop and validate our models and update estimates and projections using richer, more real time real-world data.

## Conclusions

If the relative impact of COVID-19 on mortalty over 1 year were 20% (ie. same magnitude as the winter vs summer mortality excess) then the number of excess deaths would be 0 when 1 in 100 000, 14861 when 1 in 10 and 118885 when 8 in 10 are infected. However, if the relative impact is double the mortality risk, then there would be 7, 68957 and 551659 deaths at corresponding levels of infection. These results may inform targeting those at highest risk for a range of preventive interventions.

## Data Availability

CPRD data used in this analysis can be applied for via https://www.cprd.com/research-applications
We have made online calculators available for estimation of excess mortality related to COVID-19.

https://covid19-phenomics.org/covid-excess-deaths.html

## Funding acknowledgements

AB is supported by research funding from NIHR, British Medical Association, Astra-Zeneca and UK Research and Innovation. BW and HH are National Institute for Health Research (NIHR) Senior Investigators and are funded by the National Institute for Health Research University College London Hospitals Biomedical Research Centre‥ HH work is supported by: 1. Health Data Research UK (grant No. LOND1), which is funded by the UK Medical Research Council, Engineering and Physical Sciences Research Council, Economic and Social Research Council, Department of Health and Social Care (England), Chief Scientist Office of the Scottish Government Health and Social Care Directorates, Health and Social Care Research and Development Division (Welsh Government), Public Health Agency (Northern Ireland), British Heart Foundation and Wellcome Trust. 2. The BigData@Heart Consortium, funded by the Innovative Medicines Initiative-2 Joint Undertaking under grant agreement No. 116074. This Joint Undertaking receives support from the European Union’s Horizon 2020 research and innovation programme and EFPIA; it is chaired, by DE Grobbee and SD Anker, partnering with 20 academic and industry partners and ESC. This work was supported by a National Institute of Health Research (NIHR) Clinician Scientist award (CS-2016-007) to L.S.

---

**Suppl Table 1.** High-risk mortality groups for COVID-19 infection in 3,862,012 individuals by age and gender.

|  | Men<br>Age ≤70 years<br>N=1696728 | Men<br>Age>70 years<br>N=207349 | Women<br>Age≤70 years<br>N=1634552 | Women<br>Age>70 years<br>N=323383 |
| --- | --- | --- | --- | --- |
| **Demographics** |  |  |  |  |
| Age (years), mean (SD) | 43.5 (11.66) | 78.1 (6.11) | 43.5 (11.91) | 80.2 (7.03) |
| IMD quintile |  |  |  |  |
| 1 (Least deprived) | 384993 (22.7) | 45735 (22.1) | 382183 (23.4) | 69913 (21.6) |
| 2 | 368204 (21.7) | 47890 (23.1) | 362914 (22.2) | 74205 (22.9) |
| 3 | 353908 (20.9) | 45247 (21.8) | 344154 (21.1) | 71130 (22) |
| 4 | 317818 (18.7) | 37921 (18.3) | 298922 (18.3) | 59482 (18.4) |
| 5 (Most deprived) | 270037 (15.9) | 30326 (14.6) | 244771 (15) | 48276 (14.9) |
| Practice region |  |  |  |  |
| East Midlands | 54965 (3.2) | 6669 (3.2) | 51993 (3.2) | 9924 (3.1) |
| East Of England | 196981 (11.6) | 23810 (11.5) | 188834 (11.6) | 36366 (11.2) |
| London | 297576 (17.5) | 27188 (13.1) | 289471 (17.7) | 43033 (13.3) |
| North East | 33062 (1.9) | 4134 (2) | 31491 (1.9) | 6241 (1.9) |
| North West | 230831 (13.6) | 26459 (12.8) | 217496 (13.3) | 42577 (13.2) |
| South Central | 218027 (12.8) | 26705 (12.9) | 210615 (12.9) | 42158 (13) |
| South East Coast | 213015 (12.6) | 28402 (13.7) | 208722 (12.8) | 43896 (13.6) |
| South West | 203035 (12) | 31265 (15.1) | 197035 (12.1) | 47706 (14.8) |
| West Midlands | 183652 (10.8) | 23857 (11.5) | 173924 (10.6) | 37433 (11.6) |
| Yorkshire & The Humber | 65584 (3.9) | 8860 (4.3) | 64971 (4) | 14049 (4.3) |
| **Risk factors recommended for social isolation** |  |  |  |  |
| CVD | 45650 (2.7) | 38158 (18.4) | 29668 (1.8) | 45593 (14.1) |
| Diabetes (any type) | 42901 (2.5) | 17928 (8.6) | 29791 (1.8) | 21048 (6.5) |
| Chronic kidney disease | 8202 (0.5) | 15036 (7.3) | 10136 (0.6) | 26023 (8) |
| COPD | 8990 (0.5) | 9763 (4.7) | 7475 (0.5) | 8778 (2.7) |
| BMI>40* | 8241 (1.9) | 443 (0.7) | 21042 (3.7) | 1447 (1.6) |
| Corticosteroid rx | 28935 (1.7) | 12215 (5.9) | 43894 (2.7) | 20934 (6.5) |
| N risk factors |  |  |  |  |
| 0 | 1574734 (92.8) | 138916 (67) | 1512639 (92.5) | 229571 (71) |
| 1 | 104205 (6.1) | 48663 (23.5) | 104972 (6.4) | 69704 (21.6) |
| 2 | 15049 (0.9) | 15217 (7.3) | 14230 (0.9) | 19082 (5.9) |
| ≥3 | 2740 (0.2) | 4553 (2.2) | 2711 (0.2) | 5026 (1.6) |
| **Other risk factors** |  |  |  |  |
| Hypertension | 134650 (7.9) | 66955 (32.3) | 113473 (6.9) | 101135 (31.3) |
| ACE inhibitor rx | 73513 (4.3) | 38847 (18.7) | 50299 (3.1) | 52263 (16.2) |
| NSAID rx | 204502 (12.1) | 40412 (19.5) | 262563 (16.1) | 71611 (22.1) |

\*BMI data available for 425782, 65549, 567034 and 89631 patients in the respective categories

**Suppl Table 2:** High-risk mortality groups for COVID-19 infection in 3,862,012 individuals by underlying cardiovascular disease, diabetes, chronic kidney disease and chronic obstructive pulmonary disease.

|  | No CVD<br>N=3702943 | CVD<br>N=159069 | No DM<br>N=3750344 | DM<br>N=111668 | No CKD<br>N=3802615 | CKD<br>N=59397 | No COPD<br>N=3827006 | COPD<br>N=35006 |
| --- | --- | --- | --- | --- | --- | --- | --- | --- |
| **Demographics** |  |  |  |  |  |  |  |  |
| Age (years), mean (SD) | 47.5 (16.2) | 68.9 (15.1) | 48.0 (16.5) | 62.5 (15.8) | 48.0 (16.4) | 74.5 (13.9) | 48.2 (16.6) | 70.0 (12.0) |
| Women | 1882674 (50.8) | 75261 (47.3) | 1907096 (50.9) | 50839 (45.5) | 1921776 (50.5) | 36159 (60.9) | 1941682 (50.7) | 16253 (46.4) |
| IMD quintile |  |  |  |  |  |  |  |  |
| 1 (Least deprived) | 852344 (23) | 30480 (19.2) | 863864 (23) | 18960 (17) | 870934 (22.9) | 11890 (20) | 878254 (22.9) | 4570 (13.1) |
| 2 | 818130 (22.1) | 35083 (22.1) | 830392 (22.1) | 22821 (20.4) | 839787 (22.1) | 13426 (22.6) | 847030 (22.1) | 6183 (17.7) |
| 3 | 780150 (21.1) | 34289 (21.6) | 790522 (21.1) | 23917 (21.4) | 801513 (21.1) | 12926 (21.8) | 807186 (21.1) | 7253 (20.7) |
| 4 | 683280 (18.5) | 30863 (19.4) | 690516 (18.4) | 23627 (21.2) | 702791 (18.5) | 11352 (19.1) | 706051 (18.4) | 8092 (23.1) |
| 5 (Most deprived) | 565213 (15.3) | 28197 (17.7) | 571159 (15.2) | 22251 (19.9) | 583638 (15.3) | 9772 (16.5) | 584546 (15.3) | 8864 (25.3) |
| Practice region |  |  |  |  |  |  |  |  |
| East Midlands | 117034 (3.2) | 6517 (4.1) | 120284 (3.2) | 3267 (2.9) | 122424 (3.2) | 1127 (1.9) | 122539 (3.2) | 1012 (2.9) |
| East Of England | 428720 (11.6) | 17271 (10.9) | 434154 (11.6) | 11837 (10.6) | 440118 (11.6) | 5873 (9.9) | 442772 (11.6) | 3219 (9.2) |
| London | 638219 (17.2) | 19049 (12) | 635511 (16.9) | 21757 (19.5) | 645274 (17) | 11994 (20.2) | 652256 (17) | 5012 (14.3) |
| North East | 70863 (1.9) | 4065 (2.6) | 73103 (1.9) | 1825 (1.6) | 74157 (2) | 771 (1.3) | 73814 (1.9) | 1114 (3.2) |
| North West | 491655 (13.3) | 25708 (16.2) | 502994 (13.4) | 14369 (12.9) | 509907 (13.4) | 7456 (12.6) | 510965 (13.4) | 6398 (18.3) |
| South Central | 478376 (12.9) | 19129 (12) | 483848 (12.9) | 13657 (12.2) | 490230 (12.9) | 7275 (12.2) | 493624 (12.9) | 3881 (11.1) |
| South East Coast | 475437 (12.8) | 18598 (11.7) | 478353 (12.8) | 15682 (14) | 486524 (12.8) | 7511 (12.6) | 489787 (12.8) | 4248 (12.1) |
| South West | 455024 (12.3) | 24017 (15.1) | 464293 (12.4) | 14748 (13.2) | 470063 (12.4) | 8978 (15.1) | 474405 (12.4) | 4636 (13.2) |
| West Midlands | 401024 (10.8) | 17842 (11.2) | 407566 (10.9) | 11300 (10.1) | 412622 (10.9) | 6244 (10.5) | 414886 (10.8) | 3980 (11.4) |
| Yorkshire & The Humber | 146591 (4) | 6873 (4.3) | 150238 (4) | 3226 (2.9) | 151296 (4) | 2168 (3.7) | 151958 (4) | 1506 (4.3) |
| **Risk factors recommended for social isolation** |  |  |  |  |  |  |  |  |
| CVD | 0 (0) | 159069 (100) | 139671 (3.7) | 19398 (17.4) | 142439 (3.7) | 16630 (28) | 149642 (3.9) | 9427 (26.9) |
| Diabetes (any type) | 92270 (2.5) | 19398 (12.2) | 0 (0) | 111668 (100) | 98536 (2.6) | 13132 (22.1) | 107908 (2.8) | 3760 (10.7) |
| Chronic kidney disease | 42767 (1.2) | 16630 (10.5) | 46265 (1.2) | 13132 (11.8) | 0 (0) | 59397 (100) | 56039 (1.5) | 3358 (9.6) |
| COPD | 25579 (0.7) | 9427 (5.9) | 31246 (0.8) | 3760 (3.4) | 31648 (0.8) | 3358 (5.7) | 0 (0) | 35006 (100) |
| BMI>40* | 28849 (2.7) | 2324 (2.9) | 25456 (2.4) | 5717 (6.9) | 30006 (2.7) | 1167 (3.4) | 30583 (2.7) | 590 (3.3) |
| Corticosteroid rx | 93768 (2.5) | 12210 (7.7) | 99573 (2.7) | 6405 (5.7) | 100144 (2.6) | 5834 (9.8) | 91539 (2.4) | 14439 (41.2) |
| N risk factors |  |  |  |  |  |  |  |  |
| 0 | 3455860 (93.3) | 0 (0) | 3455860 (92.1) | 0 (0) | 3455860 (90.9) | 0 (0) | 3455860 (90.3) | 0 (0) |
| 1 | 214287 (5.8) | 113257 (71.2) | 252996 (6.7) | 74548 (66.8) | 297877 (7.8) | 29667 (49.9) | 314831 (8.2) | 12713 (36.3) |
| 2 | 29714 (0.8) | 33864 (21.3) | 35760 (1) | 27818 (24.9) | 42380 (1.1) | 21198 (35.7) | 48624 (1.3) | 14954 (42.7) |
| ≥3 | 3082 (0.1) | 11948 (7.5) | 5728 (0.2) | 9302 (8.3) | 6498 (0.2) | 8532 (14.4) | 7691 (0.2) | 7339 (21) |
| **Other risk factors** |  |  |  |  |  |  |  |  |
| Hypertension | 332751 (9) | 83462 (52.5) | 355583 (9.5) | 60630 (54.3) | 378532 (10) | 37681 (63.4) | 402966 (10.5) | 13247 (37.8) |
| ACE inhibitor rx | 162700 (4.4) | 52222 (32.8) | 172910 (4.6) | 42012 (37.6) | 191583 (5) | 23339 (39.3) | 207045 (5.4) | 7877 (22.5) |
| NSAID rx | 542395 (14.6) | 36693 (23.1) | 553123 (14.7) | 25965 (23.3) | 565096 (14.9) | 13992 (23.6) | 571656 (14.9) | 7432 (21.2) |

\*BMI data available for 1067854 patients with no CVD, 80142 patients with CVD, 1064775 with no DM, 83221 with DM, 1113396 with no CKD, 34600 with CKD, 1130189 with no COPD and 17807 with COPD
*COPD: Chronic obstructive pulmonary disease, CKD: Chronic kidney disease, CVD: cardiovascular disease, DM: diabetes mellitus*

